# The VPS35 p.A320V variant segregates with Parkinson’s disease in a pesticide-exposed family

**DOI:** 10.64898/2026.08.21.26358613

**Authors:** Sophie Glendinning, José M. Arbelo, Lola Díaz-Feliz, Rocío Malo de Molina Zamora, Sara Gomes, Alexia T. Sánchez-Reyes, Kimmy Su, Dorothee Cole, Frank Hsieh, Owen A. Ross, Alexandra I. Soto-Beasley, Zbigniew K. Wszolek, Han-Joon Kim, Jung Hwan Shin, Shen-Yang Lim, Ai Huey Tan, Azlina Ahmad-Annuar, Yi Wen Tay, Teresa Kleinz, Christine Klein, Dario Alessi, Alexander Zimprich, Pau Pastor, Esther Sammler, Global Parkinson’s Genetics Program (GP2), Veterans Parkinson’s Disease Genetics Initiative, Cyrus P. Zabetian, Oswaldo Lorenzo-Betancor

**Author notes:** Correspondence to: Oswaldo Lorenzo-Betancor, Research and Development (S-182), VA Puget Sound Healthcare System, 1660 S. Columbian Way, Seattle, WA 98108. A complete list of members of the Veterans Parkinson’s Disease Genetics Initiative is provided in the Supplementary Material.

## Abstract

**Background:** The VPS35 p.D620N variant causes autosomal dominant Parkinson’s disease (PD) and has been shown to activate the LRRK2 kinase pathway, resulting in increased Rab substrate phosphorylation in peripheral immune cells and elevated urinary bis(monoacylglycero)phosphate (BMP) levels. Recently, a *VPS35* variant of unknown significance (c.959C>T; p.A320V) was described in two late-onset sporadic PD patients.

**Methods:** We ascertained a family from the Canary Islands in which six siblings were chronically exposed to high doses of pesticides. Three siblings developed levodopa-responsive, akinetic-rigid PD, while the other three remained unaffected. Whole-exome sequencing was performed in the three affected siblings. The frequency of the resulting candidate variant was assessed in 23,327 PD patients and 9,235 controls from four independent cohorts. Members of this pedigree and unrelated controls were assessed for LRRK2 kinase activity in monocytes and neutrophils and BMP levels in urine.

**Results:** The three affected siblings were all heterozygous for p.A320V, whereas the three unaffected siblings did not carry the variant. In the combined PD case-control cohort, p.A320V was identified in six patients and one control. However, unlike p.D620N, heterozygous carrier status for p.A320V was not associated with increased LRRK2 kinase activity or elevated urine BMP levels.

**Conclusions:** While VPS35 p.A320V co-segregated with PD in this family, it did not exhibit the characteristic LRRK2-associated biomarker signature observed in VPS35 p.D620N carriers. It is possible that p.A320V exerts a subtle effect on VPS35 function that was not captured by the assays performed and that chronic pesticide exposure contributed to disease penetrance in this pedigree.

## 1. Introduction

Pathogenic variants in the *vacuolar protein sorting 35* (*VPS35*) gene are a rare cause of autosomal dominant Parkinson’s disease (PD). The first pathogenic variant in *VPS35* (c.1858G>A; p.D620N) was initially described in 2011 in two independent families from central Europe.^1, 2^ A total of 63 PD patients have been reported to carry p.D620N,^1–14^ and most had a late age at onset (AAO; average=50.6, SD=8.8, range=34-68). In addition, 19 rare missense *VPS35* variants of unknown significance have been reported in the gene in another 30 PD patients.^1, 2, 6, 8, 10, 15–20^ This includes p.A320V (c.959C>T), which was described in two individuals with late-onset sporadic PD.^18^ This variant is currently classified as either of “uncertain significance” or “likely benign” in the *ClinVar* database (https://www.ncbi.nlm.nih.gov/clinvar/). Importantly, VPS35 p.D620N has been shown to activate the LRRK2 kinase pathway^21^, resulting in increased Rab substrate phosphorylation in peripheral immune cells and elevated urinary levels of bis(monoacylglycerol)phosphate (BMP) phospholipid isomer in heterozygous variant carriers^22^ Here, we report a family with akinetic-rigid PD in which affected members carry VPS35 p.A320V and present results of assays assessing LRRK2 kinase activity and urine BMP measurements.

## 2. Methods

### 2.1 Study Participants

Six siblings (three affected and three healthy) from the Canary Islands and five additional relatives were recruited at the University of Navarra School of Medicine and the University Fernando Pessoa-Canarias, respectively (Figure 1A). All were examined by a movement disorder specialist (JMA and RMMZ) and had a blood sample drawn for DNA extraction. The DNA samples from the parents (II.1 and II.2) were not available.

**Figure 1.**
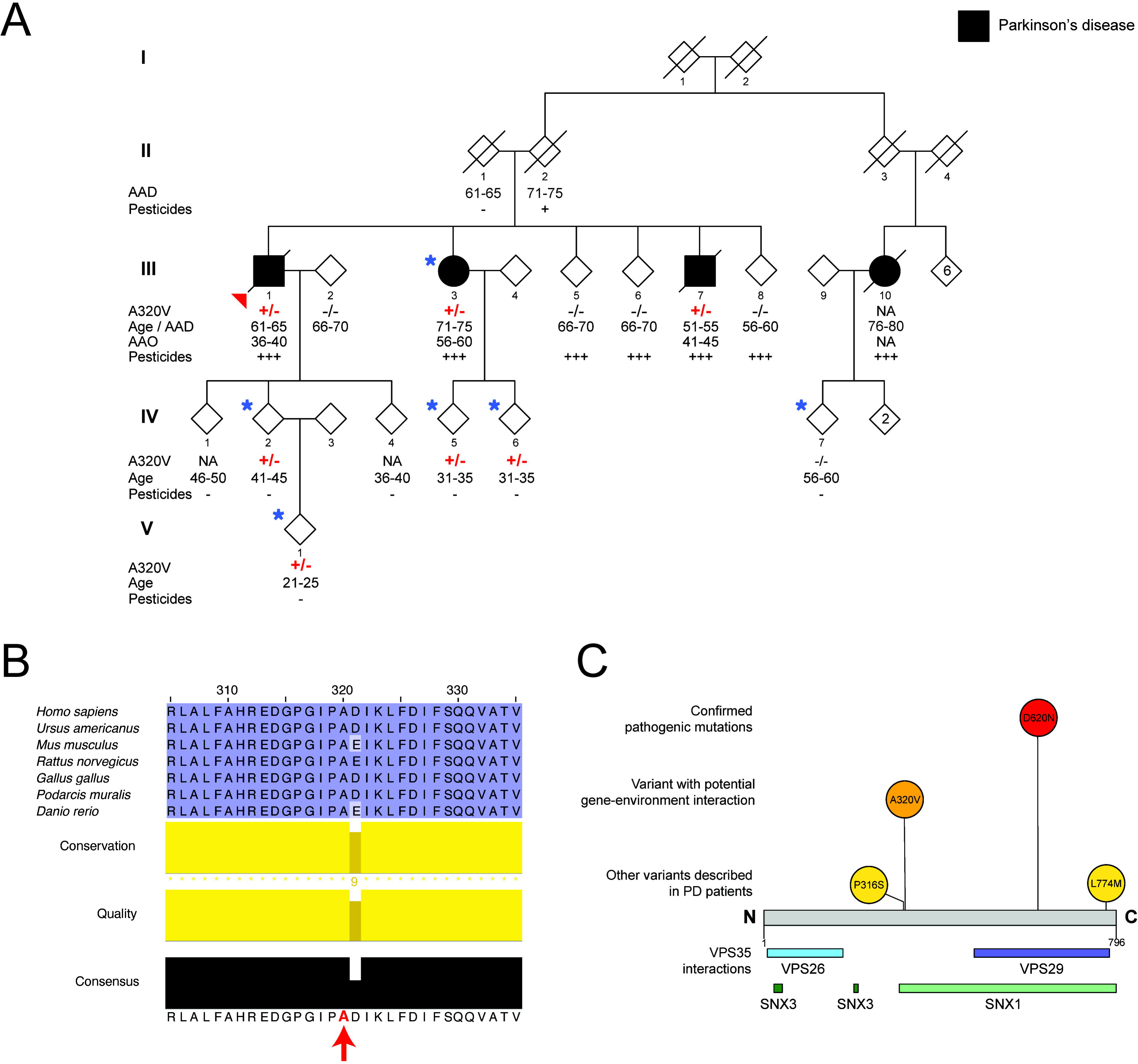
A. Simplified pedigree. Arrowhead = proband; AAD = age at death; AAO = age at onset; +/- = mutation carrier; -/- = reference allele carrier; +++ = high pesticide exposure; + = low pesticide exposure; - = no pesticide exposure; * = individuals for which neutrophils, monocytes, and urine samples were available. **B. VPS35 p.A320 amino acid conservation across species.** Red arrow = location of the p.A320 amino acid. **C. Schematic representation of the VPS35 protein**. The upper panel shows pathogenic mutations and possible pathogenic mutations described to date. p.P316S has been described in two cases and a healthy individual from the same family.^1^ p.L774M has been described in six cases and one healthy control.^6^ The lower panel shows retromer protein interaction regions.

For functional analysis of pT73-Rab10 levels, fresh blood samples were collected from affected family members (III.3, IV.2, IV.5, IV.6, IV.7, and V.1) at the University Fernando Pessoa Canarias, and from four unrelated non-carrier controls (D01–D04) at the University of Dundee. For analysis of urinary BMP levels, fresh urine samples were obtained from the same participants. In addition, six p.D620N-VPS35 carriers (D05–D10) were recruited at the University of Lübeck for urinary BMP analysis. A summary of the participants included in the functional analyses is provided in Supplementary Table 1.

**Table 1.** Clinical phenotype of individuals with PD.

| Individual | Sex | Pesticide exposure | AAO | Age at last assessment | Initial symptom | Resting tremor | Rigidity | Bradykinesia | Postural instability | Cognitive impairment | L-dopa response | Motor fluctuations | Dyskinesias | DRT-induced complications | Presynaptic dopaminergic deficit (DAT-SPECT) | Other findings |
| --- | --- | --- | --- | --- | --- | --- | --- | --- | --- | --- | --- | --- | --- | --- | --- | --- |
| III.1 | M | Daily / work-related | 36-40 | 61-65 | Rigidity and rest tremor of right hand | + | + | + | Yes | - | + | + | + | RBD | NA | DBS-induced neuropsychiatric manifestations |
| III.3 | F | Daily / work-related | 56-60 | 66-70 | Right shoulder pain and arm rigidity | + | + | + | No | - | + | + | + | “off” dystonia | NA | None |
| III.7 | M | Daily / Work-related | 41-45 | 51-55 | Fatigue and pain of right upper extremity | - | + | + | No | + | + | + | + | “off” dystonia | Yes | sleep apnea |

AAO = age at onset; DAT-SPECT = dopamine transporter (DAT) - single photon emission computed tomography <sup>35</sup>; DBS = deep brain stimulation; DRT = dopamine replacement therapy; F = female; M = male; NA = Not available; RBD = rapid eye movement (REM) sleep behavior disorder.

The study was approved by the Institutional Review Boards of the participating institutions, and all participants provided written informed consent prior to enrollment and sample collection. All procedures were conducted in accordance with the principles of the Declaration of Helsinki.

### 2.2 Genetic analyses

Genomic DNA was isolated from the peripheral blood of available participants. The proband was screened for copy number variation in known PD genes using multiplex ligation-dependent probe amplification (MLPA; MRC-Holland, Amsterdam, Netherlands). Exome sequencing was performed with the Sure Select V4 + UTR exome capture kit (Agilent Technologies, Santa Clara, CA, USA) using 2.1 µg of genomic DNA from each subject following the manufacturer’s standard protocol on a HiSeq 2000 sequencer (Illumina, San Diego, CA, USA). We performed alignment and base calling using custom in-house-built pipelines. SNP annotation was performed using ANNOVAR.^23^ Variants were numbered according to standard nomenclature (http://www.hgvs.org/mutnomen/)^24^ based on RefSeq NM_018206.6. and NP_060676.2 accession numbers.

### 2.3. Variant selection criteria

Analysis and variant prioritization were performed using an in-house-built SQL pipeline. Low-quality variants (genotype quality <20 or read depth <10) were excluded from the analysis. Variants with a minor allele frequency (MAF) >0.1% in the Genome Aggregation Database (gnomAD) v.4.1.0 were excluded. Variants that were not either missense mutations, coding deletions or insertions, frameshift mutations, intronic or synonymous variants located near splice sites (≤5 nucleotides) were further removed from the analysis and only variants with a Combined Annotation Dependent Depletion (CADD) score >20 were retained (Supplementary Table 2). Variants located in known PD genes were prioritized for segregation analysis with Sanger sequencing in the available relatives.

### 2.4 Candidate variant sequencing and co-segregation

*VPS35* c.959C>T (p.A320V) was Sanger-sequenced in all individuals to confirm a true call and to check co-segregation with disease. Primer pairs were designed using Primer3.^25^

### 2.5 Variant screening

The *VPS35* c.959C>T (p.A320V) variant was screened in 567 independent sporadic and familial individuals with PD from mainland Spain, and 141 independent sporadic and familial individuals with PD and 63 healthy controls from the Parkinson’s disease Genetics in the Canary Islands (PDGenCI) cohort with a TaqMan assay using 1ul DNA. Primers and conditions are available on request. In addition, we queried whole genome sequencing (WGS) datasets from the Global Parkinson’s Genetics Program (GP2; 21,206 PD participants and 9,172 controls) and the Veterans Parkinson’s Disease Genetics Initiative (Vet-PD) (1,413 PD participants) to screen for additional VPS35 p.A320 carriers.

### 2.6 Neutrophil and monocyte isolation, treatment, and lysis

Neutrophils and monocytes were isolated from peripheral human blood using immunomagnetic negative selection in six members of the family. Neutrophils were isolated from 10 mL of blood using the EasySep™ Human Neutrophil Isolation Kit (STEMCELL Technologies, Cat# 19666), and monocytes were isolated from 20 mL of blood using the EasySep™ Human Monocyte Isolation Kit (STEMCELL Technologies, Cat# 19359), as described previously.^21, 26^

Live, isolated neutrophils and monocytes were pelleted and resuspended in RPMI media (Thermo Fisher). The cell suspensions were divided into two tubes and incubated with either 200 nM MLi-2 or vehicle control (DMSO) for 15 min at room temperature (RT). Following incubation, cells were pelleted and resuspended in ice-cold lysis buffer (50 mM Tris/HCl pH 7.5, 1% Triton X-100, 1 mM EGTA, 1 mM sodium orthovanadate, 50 mM sodium fluoride, 0.1% 2-mercaptoethanol, 10 mM 2-glycerophosphate, 5 mM sodium pyrophosphate, 1 μg/ml mycrocystin-LR (Enzo Life Sciences), 270 mM sucrose, 0.5 mM diisopropylfluorophosphate (DIFP) (Sigma, Cat# D0879) supplemented with Complete EDTA-free protease inhibitor cocktail (Roche, Cat# 11836170001)). Cell lysates were snap-frozen and stored at -80°C.

### 2.7 Immunoblotting

Cell lysates were clarified at 17,000 x g for 20 mins at 4°C, and the protein concentration of the resulting supernatant was measured using the Pierce BCA Protein Assay Kit (Thermo Fisher). Lysates were mixed with the NuPAGE LDS Sample Buffer (Life Technologies) supplemented with 5% (v/v) 2-mercaptoethanol, then boiled at 90°C for 10 minutes. Twenty μg of protein was loaded onto NuPAGE 4–12% Bis-Tris Midi Gels (Thermo Fisher) and electrophoresed in NuPAGE MOPs SDS running buffer (Thermo Fisher). Protein transfer onto nitrocellulose membranes (GE Healthcare, Amersham Protran Supported 0.45 μm NC) was performed on ice, with 90 V for 90 mins in transfer buffer (48 mM Tris–HCl and 39 mM glycine supplemented with 20% methanol). Membranes were blocked for 60 mins at RT with 5% skim milk dissolved in TBS-T (20 mM Tris–HCl, pH 7.5, 150 mM NaCl, and 0.1% (v/v) Tween 20). Membranes were then washed once before an overnight incubation at 4°C with primary antibodies diluted in 5% BSA dissolved in TBS-T. After overnight incubation, membranes were washed for 5 minutes at RT 3x with TBS-T before incubation with secondary antibodies diluted in 5% skim milk for 1 hour at RT. Membranes were then washed for 10 mins at RT three times before image acquisition with the Odyssey CLx imaging system (LI-COR Biosciences). Quantification of protein bands was performed using Image Studio Software (LI-COR Biosciences).

### 2.8 Antibodies

Mouse monoclonal anti-total LRRK2 mouse (NeuroMab #75-253) and rabbit monoclonal anti-pS935 LRRK2 (ab133450) were used at a final concentration of 1 μg/mL and were purified in-house as described previously.^27^ Mouse monoclonal anti-total Rab10 (0680-100, Nanotools) and rabbit monoclonal anti-pT73-Rab10 (ab241060, Abcam) were used at a final concentration of 1 μg/mL. Mouse monoclonal anti-GAPDH (sc-32233, Santa Cruz) was used at a concentration of 50 ng/mL. Mouse monoclonal anti-VPS35 (SMC-602, StressMarq) and rabbit monoclonal anti-VPS26 (ab181352, Abcam) were used at a 1:2000 dilution. The secondary antibodies, anti-rabbit (Licor, 926-32213) and anti-mouse (Licor, 926-68072), were used at a 1:20,000 dilution.

### 2.9 Targeted mass spectrometry

One hundred μg of cleared neutrophil lysate and 50μg of cleared monocyte lysate were prepared and processed for targeted mass spectrometry of pT73-Rab10 and Total Rab10 as previously described.^28^ Proteins were reduced, alkylated, and digested using a single-pot SP3 trypsin/Lys-C workflow, after which peptides were acidified and aliquots reserved for total Rab10 measurements. PT73-Rab10 peptides were quantified by automated immunoprecipitation using Protein G magnetic beads conjugated with specific antibodies, with heavy-labeled peptide standards added during peptide resuspension.^28^ Enriched peptides were eluted with trifluoroacetic acid. Total Rab10 peptides were measured following resuspension in LC buffer containing labeled peptide standards.^28^

Peptides were analyzed by parallel reaction monitoring (PRM) on an EvoSep One liquid chromatography system coupled to an Orbitrap Exploris 480 mass spectrometer (Thermo Fisher Scientific), using predefined precursor isolation windows and optimized collision energies. PRM data were analyzed in Skyline (version 24.1.0.199) using light-to-heavy peptide ratios for quantification. External calibrations were used to determine absolute peptide concentrations.

### 2.10 Urine BMP analysis

Urine was collected from five VPS35 p.A320V carriers: one PD patient and four non-manifesting carriers (NMC). In addition, urine samples from six *VPS35* p.620N carriers (four PD patients and two NMCs) and five non-mutant carriers were used as positive and negative controls, respectively. One of these negative controls was a wild-type relative. Urine processing for BMP analysis was performed as previously described.^22^ Briefly, midstream urine samples were collected from participants and processed within 30 minutes of collection. Samples were centrifuged at 2,500 × g for 15 minutes at 4°C to remove sediment. The supernatant was aliquoted into labeled Eppendorf tubes, snap-frozen, and stored at -80°C. Samples were shipped on dry ice and maintained at -80°C until further processing by Nextcea, Inc. (Woburn, MA). Simultaneous quantification of BMP species was performed by Nextcea, Inc., with analysts blinded to participant identity and clinical status, using a multiplexed UPLC-MS/MS method as described earlier.^29^ Sample normalization, calibration, and data processing were conducted by Nextcea, Inc. according to the same methodology.

## 3. Results

### 3.1 Clinical findings

#### Individual III.1

The proband, a male enrolled in his early 60’s, passed away after a long history of PD in his early 60’s (Figure 1A; individual III.1). He drank well water and worked in agricultural greenhouses for several decades with high exposure to pesticides without protective equipment. In his early 20’s, he was hospitalized because of an acute pesticide intoxication. In his late 30’s, he complained of rigidity and tremor in his right hand. He was started on levodopa with initial clinical improvement. He later developed wearing-off, severe dyskinesias, and non-motor complications such as painful off periods, constipation, and REM sleep behavior disorder (RBD). In his early 50’s, he underwent deep-brain stimulation (DBS) surgery. After the procedure, the patient showed permanent behavioral changes, aggressiveness, irritability, and worsening of motor symptoms. Subsequently, his DBS device was turned off. He was started on carbidopa/levodopa intraduodenal gel with significant behavioral improvement and moderate motor improvement.

#### Individual III.3

The proband’s sister did not display signs of parkinsonism at initial enrollment. She worked in greenhouses for several decades and was exposed to high levels of pesticides. However, her motor symptoms did not begin until her early 60’s, when she complained of right shoulder pain and arm rigidity (Figure 1A; individual III.3). She was started on levodopa with a good initial clinical response. Her motor symptoms worsened, and she developed marked motor fluctuations with “off” dystonia and continuous dyskinesias during “on” periods. In her late 60’s, she was started on a continuous subcutaneous apomorphine pump, which provided a slight improvement of her motor fluctuations.

#### Individual III.7

The proband’s brother (Figure 1A; individual III.7) was in his late 40’s at enrollment. He began working in agricultural greenhouses when he was a teenager. In his early 40’s, he complained of tiredness, extreme fatigue, and pain in his right upper extremity. He later experienced motor clumsiness, bradykinesia, and difficulties with handwriting, but he never developed tremor. He was diagnosed with PD in his early 40’s and was initially started on a dopamine agonist. Levodopa was subsequently added, and he rapidly developed the wearing-off phenomenon, with freezing of gait and “off” dystonia. When he was in his late 40’s, he underwent a neuropsychological battery that showed concentration difficulties, visuospatial impairment, minor deficits in recent memory, and diminished verbal fluency. When he was in his early 50’s, he developed focal seizures. An EEG showed interhemispheric asymmetry. He passed away in his early 50’s due to a grade 4 glioblastoma.

### 3.2 Genetic findings

Exome sequencing revealed 38 variants fulfilling selection criteria (Supplementary Table 2). One of them was in the *VPS35* gene (c.959C>T, p.A320V), making it the most plausible cause of disease. The missense variant was present in the three affected members and absent in the three healthy siblings (Figure 1A). The allele frequency was very low (0.9 x 10^-4^) among 730,947 exomes and 76,215 genomes available from the *Genome Aggregation Database* v4.1.0. The variant is located between VPS35 α-helices 14 and 15,^30^ is evolutionarily conserved across the known vertebrate homologs of the VPS35 protein (Figure 1B) and is predicted to be deleterious based on its high CADD score (23.3). No plausible pathogenic coding or structural variants that met our predefined criteria were identified in any other PD-related genes. VPS35 p.A320V was not observed in a cohort of 567 idiopathic and familial PD cases from mainland Spain, nor in a cohort of 141 PD cases and 63 controls from the PDGenCI cohort.

The screening of the WGS datasets from the GP2 and Vet-PD cohorts showed an overrepresentation of VPS35 p.A320V in cases (6/22,619; MAF = 2.65 x 10^-4^) compared to controls (1/9,172; MAF = 1.09 x 10^-4^) that did not reach significance (p-value = 0.40) (carrier details in Supplementary Table 3). The demographics for all PD cases and controls are in Supplementary Tables 4 and 5, respectively.

### 3.3 Functional analyses

LRRK2 phosphorylates a subset of Rab-GTPases, including Rab10,^31^ making Rab phosphorylation a well-established biomarker of LRRK2 kinase pathway activity.^21, 32, 33^ To investigate whether the VPS35 p.A320V mutation affected LRRK2 kinase activity, we isolated neutrophils and monocytes from fresh blood from six members of the affected family (III.3, IV.2, IV.5, IV.6, IV.7, and V.1) and four unrelated non-carrier control donors. Among the family members analyzed, five were heterozygous VPS35 p.A320V carriers, including one individual with PD and four non-manifesting carriers (NMC). LRRK2 kinase pathway activity was assessed using immunoblotting and targeted mass spectrometry of pT73-Rab10, normalized against total Rab10 levels (Fig. 2). Analysis by immunoblotting revealed no significant overall increase in pT73-Rab10 levels in p.A320V carriers relative to non-carriers, neither in neutrophils (Fig. 2C) nor in monocytes (Fig. 2E). Similarly, targeted mass-spectrometry did not identify significant differences in pT73-Rab10 levels between the groups. Treatment with the selective LRRK2 kinase inhibitor MLi-2 significantly reduced pT73-Rab10 levels in both neutrophils and monocytes (Fig. 2), confirming assay responsiveness and LRRK2 dependency of the signal (neutrophils: non-carriers p = 0.03, p.A320V carriers p = 0.02; monocytes: non-carriers p = 0.01, p.A320V carriers p = 0.05)

**Figure 2.**
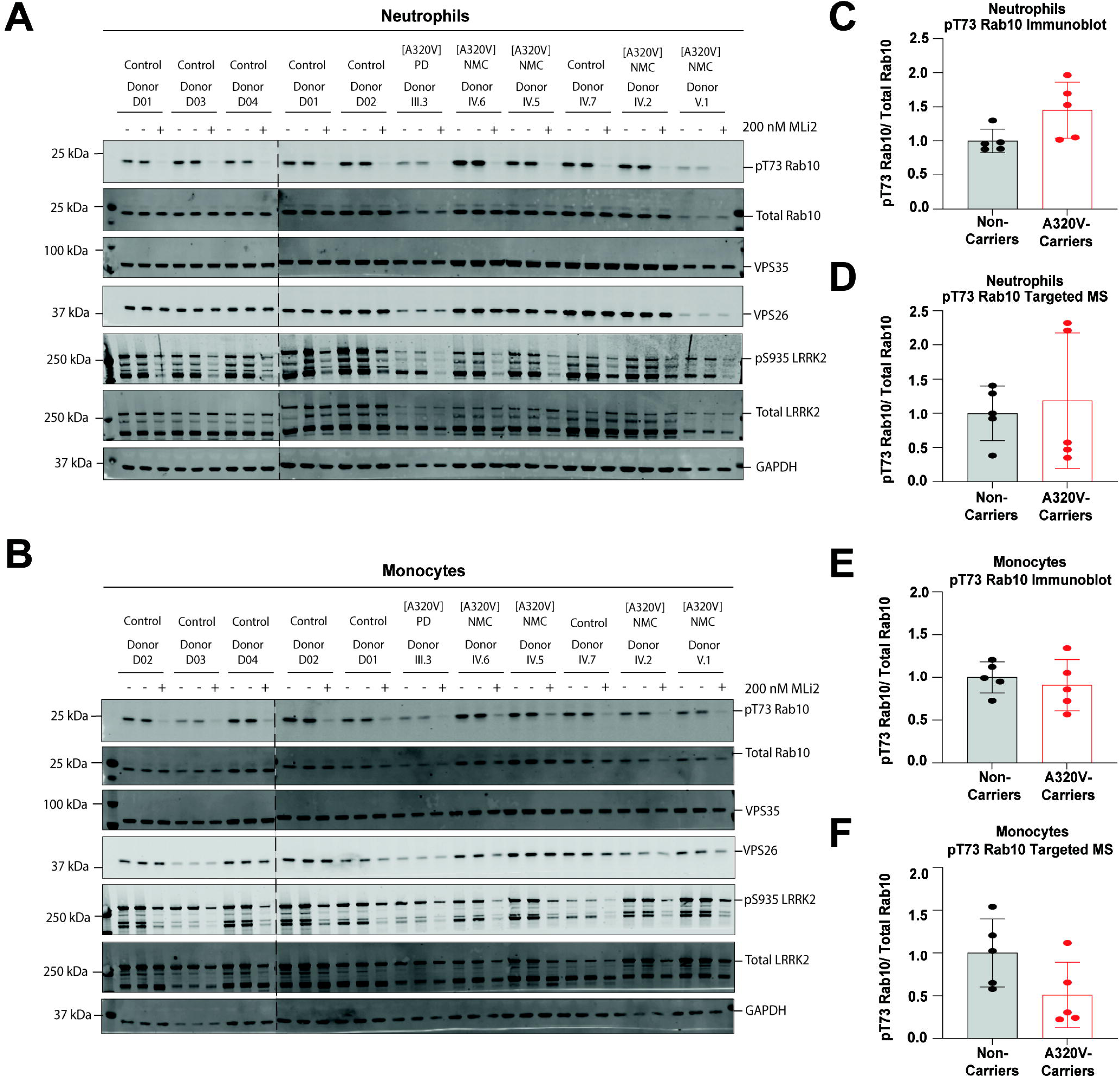
Investigating LRRK2-mediated Rab10 phosphorylation in neutrophils and monocytes from A320V mutation carriers and controls. A) Summary of donor participants; neutrophils and monocytes were isolated from six members of the affected family, including 1 VPS35 p.A320V carrier with PD (individual III.3), four NMC VPS35 p.A320V (individuals IV.2, IV.5, IV.6, and V.1), and one non-variant carrier (individual IV.7). An additional four un-related, non-mutation carriers were also included as controls (individuals D01, D02, D03, and D04). A-B) Immunoblot analysis was performed with 20 μg of protein from neutrophil and monocyte samples. Neutrophils and monocytes were treated with a LRRK2-specific small-molecule inhibitor, MLi2 (200 nM), or a vehicle control for 30 minutes prior to lysis. C-F) Quantification of LRRK2 activity with neutrophils and monocytes by immunoblot analysis and targeted mass spectrometry (MS). PT73-Rab10 levels / total Rab10 levels were normalized to the average of non-carrier (vehicle-treated control) samples. LRRK2 activity was reported as fold-change relative to the average of non-carrier (vehicle-treated controls) samples. Data were analy**z**ed using Welch’s t-test. pT73-Rab10 levels were not significantly elevated in neutrophils or monocytes collected from A320V mutation carriers (n=5) compared to non-mutation carriers (n=5).

Next, we compared urinary BMP levels among non-carriers, VPS35 p.A320V carriers, and VPS35 p.D620N carriers. Both the p.A320V and p.D620N carrier groups included individuals with PD as well as NMC. Consistent with previous findings,^22^ p.D620N carriers displayed the characteristic increase in urinary di-22:6 and di 18:1 BMP levels, whereas VPS35 p.A320V carriers showed no significant increase in either BMP isoform compared to non-carriers (Supplementary Fig. 1).

## 4. Discussion

This study is the first to demonstrate that *VPS35* c.959C>T (p.A320V) co-segregates with disease in a PD pedigree. Two of the three affected siblings had an AAO that is earlier than most patients reported with *VPS35*-related PD (III.1, 36-40 years; III.7, 41-45 years). Only individuals in generation III were exposed to high levels of pesticides, and only p.A320V carriers in this generation developed PD (Figure 1A). These observations raise the possibility that pesticide exposure influenced penetrance and possibly the AAO associated with p.A320V.

To date, p.D620N is the only well-established pathogenic VPS35 variant. Previous studies have shown that p.D620N activates the LRRK2 kinase pathway, resulting in increased Rab10 phosphorylation in peripheral immune cells from heterozygous carriers and VPS35 p.D620N knock-in mouse models,^21^ as well as elevated urinary BMP levels.^22^ Although the precise mechanism remains incompletely understood, these findings support a functional link between VPS35 p.D620N and enhanced LRRK2 kinase activity.^21^ In contrast, our analyses using quantitative immunoblotting and targeted mass spectrometry of pT73-Rab10 did not demonstrate significant differences between VPS35 p.A320V carriers and non-carriers (Figure 2).

However, subtle increases in LRRK2 kinase activity cannot be excluded. Our analyses were limited by a small sample size, and the assays and biomarkers deployed may not capture all forms of pathogenic LRRK2 pathway activation associated with VPS35 dysfunction. For example, heterozygous LRRK2 p.G2019S carriers do not consistently show significantly elevated pT73-Rab10 levels in peripheral blood despite clear evidence of pathogenicity and elevated urinary BMP levels.^26^ In contrast, previous studies in VPS35 p.D620N carriers demonstrated increased Rab phosphorylation. It therefore remains possible that VPS35 p.A320V induces more subtle or selective alterations in Rab phosphorylation, affects other Rab substrates, or influences downstream pathways not captured by the assays used in this study. Larger cohorts and complementary functional approaches may therefore be required to fully assess the biological consequences of VPS35 p.A320V.

Previous studies also reported elevated urinary BMP phospholipid isomers, particularly di-22:6 and di-18:1, in carriers of the VPS35 p.D620N as well as *LRRK2* and *GBA1* variant carriers.^22, 34^ Consistent with these findings, urinary BMP levels were elevated in VPS35 p.D620N carriers but not in VPS35 p.A320V carriers in our cohort.

In the combined case-control cohort, VPS35 p.A320V was more frequent in individuals with PD than controls, although this did not reach significance. Given the very low allele frequency of p.A320V, substantially larger datasets would be required to determine whether this variant contributes to PD risk at the population level.

Prior in vitro evidence showed that VPS35 p.A320V alters VPS35 function. Analyses of HEK-293T cells transfected with human VPS35 p.A320V or wild-type constructs and an α-synuclein construct showed that this variant alters endosomal and lysosomal functions and affects retromer-mediated protein trafficking.^18^ However, the relevance of this finding for *VPS35*-related PD is uncertain. There are no autopsy reports from *VPS35*-related PD individuals, and it is unclear whether *VPS35* pathogenic variant carriers develop Lewy bodies.^32^

Overall, while VPS35 p.A320V co-segregated with PD in this family, we did not identify the characteristic LRRK2-associated biomarker signature observed in VPS35 p.D620N carriers. Further studies are warranted to determine whether p.A320V contributes to PD susceptibility through more subtle effects on retromer and lysosomal function and whether chronic pesticide exposure acted as an additional disease-modifying factor in this pedigree.

## Data Availability

All code generated to extract the GP2 data, and the identifiers for all software used, are available on GitHub (https://github.com/GP2code/GP2-VPS35_p.A320V) and were given a persistent identifier via Zenodo (10.5281/zenodo.22085586).

## Acknowledgments

We sincerely thank all the individuals who participated in this study. Data from the Global Parkinson’s Genetics Program (GP2), from the Veterans Parkinson’s Disease Genetics Initiative (Vet-PD), and from the LRRK2 Cohort Consortium (LCC) were used to confirm that the VPS35 p.A320V variant was present in seven participants from these consortia. We used Tier 2 data from GP2 release 11 (https://zenodo.org/records/17753486). GP2 is funded by the Aligning Science Across Parkinson’s (ASAP) initiative and implemented by The Michael J. Fox Foundation for Parkinson’s Research (https://gp2.org). For a complete list of GP2 members, see https://gp2.org. The investigators within the LRRK2 Cohort Consortium (LCC) contributed to the design and implementation of the LCC and/or provided data and/or collected biospecimens but did not necessarily participate in the analysis or writing of this report. The full list of LCC investigators can be found at https://mjff.prod.acquia-sites.com/sites/default/files/media/document/LRRK2_Cohort_Consortium_Investigators_List.pdf. T.K. and C.K. would like to acknowledge support of the Global Parkinson’s Genetics Program (GP2). The views expressed in this article are those of the authors and do not necessarily reflect the position or policy of the Department of Veterans Affairs or the United States government.

## Author Roles

(1) Research Project: A. Conceptual Design, B. Organization, C. Experiment Execution, D. Clinical Data Acquisition; (2) Statistical Analysis: A. Design, B. Execution, C. Review and Critique; (3) Manuscript Preparation: A. Writing of the First Draft, B. Writing of the Final Draft, C. Editing; (4) Resources: A. Sample provision, B. Funding acquisition.

S.G.: 1A, 1B, 1C, 2A, 2B, 3C, 4A.

J.M.A.: 1D, 3C, 4A.

L.D.-F.: 1D, 3C, 4A.

R.M.M.Z.: 1D, 3C, 4A.

S.G.: 1C, 3C.

A.T.S.R: 1D, 3C, 4A.

K.S.: 1D, 3C, 4A.

D.C.: 1D, 3C, 4A.

F.H.: 1C, 2B, 3C.

O.A.R.: 1A, 1B, 3C, 4B.

A.I.S.B.: 1B, 1C, 3C.

Z.K.W.: 1D, 3C, 4A.

H.-J.K.: 1D, 3C, 4A.

J.H.S.: 1D, 3C, 4A.

S.-Y.L.: 1D, 3C, 4A.

A.H.T.: 1D, 3C, 4A.

A.A.-A.: 1D, 3C, 4A.

Y.W.T.: 1D, 3C, 4A.

T.K.: 1D, 3C, 4A.

C.K.: 1D, 3C, 4A.

D.A.: 1A, 1B, 2C, 3C.

A.Z.: 1A, 1B, 2C, 3C, 4A.

P.P.: 1A, 1B, 3C, 4A, 4B.

E.S.: 1A, 1B, 2C, 3C, 4B.

Members from the Vet-PD consortium and the Global Parkinson’s Genetics Program (GP2): 3C, 4A.

C.P.Z.: 1A, 1B, 2C, 3C, 4A, 4B.

O.L.-B.: 1A, 1B, 1C, 1D, 2A, 2B, 3A, 3B, 4A, 4B.

