## Supplementary Materials for "The VPS35 p.A320V variant segregates with Parkinson’s disease in a pesticide-exposed family"

[a]Medical Research Council (MRC) Protein Phosphorylation and Ubiquitylation Unit, Faculty of Life Sciences, University of Dundee, Dundee, United Kingdom.
[b]Department of Neurology, University Hospital San Roque, Las Palmas de Gran Canaria, Spain.
[c]School of Medicine University Fernando Pessoa-Canarias, Las Palmas de Gran Canaria, Spain.
[d]Department of Neurology, Complejo Hospitalario Universitario Insular de Las Palmas, Las Palmas de Gran Canaria, Spain.
[e]Veterans Affairs Puget Sound Health Care System, Seattle, WA, USA.
[f]Department of Neurology, University of Washington School of Medicine, Seattle, WA, USA.
[g]Veterans Affairs Loma Linda Health Care, Loma Linda, CA, USA.
[h]Nextcea, Inc. 500 West Cummings Park, Suite 4550, Woburn, MA, USA.
[i]Department of Neuroscience, Mayo Clinic Florida, Jacksonville, FL, USA.
[j]Department of Neurology, Mayo Clinic Florida, Jacksonville, FL, USA.
[k]Department of Neurology, Seoul National University Hospital, Seoul National University College of Medicine, Seoul, Korea.
[l]Division of Neurology, Department of Medicine, Faculty of Medicine, Universiti Malaya, Kuala Lumpur, Malaysia.
[m]Department of Biomedical Science, Faculty of Medicine, Universiti Malaya, Kuala Lumpur, Malaysia
[n]Institute of Neurogenetics, University of Luebeck, Luebeck, Germany.
[o]Section for Movement Disorders, University Hospital of Schleswig-Holstein, Luebeck, Germany.
[p]Department of Neurology, Medical University, Vienna, Austria.
[q]Unit of Neurodegenerative Diseases, Department of Neurology, University Hospital Germans Trias i Pujol and the Germans Trias i Pujol Research Institute (IGTP), Badalona, Barcelona, Spain.
[r]Division of Neuroscience, Faculty of Health, University of Dundee, Dundee, United Kingdom.
[$]A complete list of members of the Veterans Parkinson's Disease Genetics Initiative is provided at the end of this Supplement

**Supplementary Table 1. Details of the participants of functional analysis**

| Donor ID | Phenotype | Sex | Age at sample collection |
|---|---|---|---|
| III.3 | A320V VPS35-PD | Female | 71-75 |
| IV.2 | A320V VPS35-NMC | Female | 31-35 |
| IV.5 | A320V VPS35-NMC | Female | 31-35 |
| IV.6 | Control | Female | 56-60 |
| IV.7 | A320V VPS35-NMC | Female | 41-45 |
| V.1 | A320V VPS35-NMC | Female | 21-25 |
| D01 | Control | Female | 46-50 |
| D02 | Control | Female | 31-35 |
| D03 | Control | Female | 26-30 |
| D04 | Control | Female | 31-35 |
| D05 | VPS35[D620N]-PD | Female | 51-55 |
| D06 | VPS35[D620N]-NMC | Female | 31-35 |
| D07 | VPS35[D620N]-NMC | Male | 51-55 |
| D08 | VPS35[D620N]-PD | Female | 51-55 |
| D09 | VPS35[D620N]-PD | Male | 71-75 |
| D10 | VPS35[D620N]-PD | Female | 66-70 |

NMC = non-manifesting carrier; PD = Parkinson's disease

### Supplementary Table 2. List of candidate variants shared by the three affected PD subjects

| Gene | Chr | Position | Ref | Alt | rs identifier | Classification | cDNA | Protein | MAF* | CADD score |
|---|---|---|---|---|---|---|---|---|---|---|
| *PDE4DIP* | 1 | 149007441 | C | T | rs782304850 | splicing | c.4651+5G>A | NA | 0.000002694 | 26.3 |
| *ABCG5* | 2 | 43823985 | C | T | rs1279466034 | missense | c.1252G>A | p.D418N | 0.000001695 | 23.6 |
| *CCDC88A* | 2 | 55322591 | T | G | rs372575449 | missense | c.3096A>C | p.E1032D | 0.00002465 | 22.9 |
| *CAMKMT* | 2 | 44362148 | A | T | rs541010080 | splicing | c.138+3A>T | NA | 0.00008194 | 21.4 |
| *ALMS1* | 2 | 73386086 | A | C | rs762635802 | missense | c.218A>C | p.D73A | 0 | 25.8 |
| *SMYD5* | 2 | 73214295 | CCTT | C | rs767636363 | nonframeshift deletion | c.30_32del | p.F11del | 0.00008051 | 22.3 |
| *ZC3H6* | 2 | 112331995 | C | G | rs1376369038 | missense | c.3077C>G | p.P1026R | 0 | 24.9 |
| *MST1* | 3 | 49684128 | A | C | rs1057319 | missense | c.2078T>G | p.I693R | 0.000005933 | 26 |
| *IMPG2* | 3 | 101228792 | C | G | NA | splicing | c.3302+5G>C | NA | 0 | 23 |
| *WFS1* | 4 | 6301441 | T | C | rs550411492 | missense | c.1646T>C | p.L549P | 0.00002966 | 22.4 |
| *EVC* | 4 | 5783693 | C | G | rs138821019 | missense | c.1705C>G | p.L569V | 0.000005085 | 23.6 |
| *NPFFR2* | 4 | 72147028 | C | T | rs201155248 | missense | c.785C>T | p.A262V | 0.0001839 | 24.2 |
| *EXOC3* | 5 | 446315 | C | T | rs759279639 | missense | c.110C>T | p.A37V | 0.00001696 | 25 |
| *CMYA5* | 5 | 79729204 | C | T | rs555997523 | missense | c.439C>T | p.R147W | 0.00004832 | 25.9 |
| *LNPEP* | 5 | 96985104 | A | C | NA | missense | c.885A>C | p.E295D | 0 | 25.5 |
| *CYFIP2* | 5 | 157319762 | G | A | NA | missense | c.1279G>A | p.V427M | 0 | 24.9 |
| *TBC1D9B* | 5 | 179893372 | C | T | rs925625024 | missense | c.673G>A | p.E225K | 0.000006779 | 25.6 |
| *FAM83B* | 6 | 54940449 | G | A | rs376811005 | missense | c.1478G>A | p.R493H | 0.00006186 | 28.4 |
| *TIAM2* | 6 | 155256766 | C | T | rs143599677 | missense | c.1526C>T | p.P509L | 0.0004593 | 24.3 |
| *ARID1B* | 6 | 157181138 | G | A | rs746567959 | missense | c.3305G>A | p.R1102Q | 0.00003135 | 29.4 |
| *NDUFB9* | 8 | 124547012 | T | G | rs757935794 | missense | c.307T>G | p.C103G | 0.00008987 | 22.4 |
| *VPS28* | 8 | 144424956 | C | T | rs781982468 | missense | c.290G>A | p.R97H | 0.00001554 | 25.2 |
| *ZMYND19* | 9 | 137587819 | C | T | rs1235562329 | missense | c.116G>A | p.R39Q | 0.000004237 | 23.3 |
| *ITIH2* | 10 | 7734981 | C | T | rs768339143 | missense | c.1847C>T | p.S616F | 0.000007627 | 25.5 |
| *EMSY* | 11 | 76458300 | T | A | rs1229341075 | missense | c.363T>A | p.N121K | 0 | 24.2 |
| *RAPGEF3* | 12 | 47743548 | T | C | rs761724079 | missense | c.1807A>G | p.K603E | 0.000006781 | 23.9 |
| *POU6F1* | 12 | 51192459 | G | C | rs1272303973 | missense | c.262C>G | p.Q88E | 0.000003390 | 22.4 |
| *SETD1B* | 12 | 121817893 | A | G | rs1018114517 | missense | c.3278A>G | p.D1093G | 0.000003494 | 23.6 |
| *MYCBP2* | 13 | 77168483 | T | C | rs141717634 | missense | c.6059A>G | p.Y2020C | 0.00009237 | 27.6 |
| *RALGAPA1* | 14 | 35549222 | T | G | NA | missense | c.5991A>C | p.R1997S | 0 | 25.4 |
| *RPUSD2* | 15 | 40574206 | T | C | rs1891209012 | missense | c.1400T>C | p.F467S | 0 | 29.2 |
| *SRRM2* | 16 | 2767952 | T | C | rs763775653 | missense | c.7424T>C | p.L2475P | 0.00003944 | 22.3 |
| *VPS35* | 16 | 46674616 | G | A | rs747944333 | missense | c.959C>T | p.A320V | 0.00006361 | 23.2 |
| *SMG6* | 17 | 2282721 | G | A | rs375743887 | missense | c.2587C>T | p.R863W | 0.00001695 | 23.6 |
| *ZNF516* | 18 | 76441490 | T | C | rs751361720 | missense | c.1565A>G | p.K522R | 0.00002469 | 25.4 |
| *CNDP2* | 18 | 74520016 | G | A | rs529520577 | missense | c.1124G>A | p.G375E | 0.00002966 | 28.6 |

*MAF = minor allele frequency for Non-Finnish European population from gnomAD v4.1.0

**Supplementary Table 3. Details of the p.A320V variant carriers**

| Phenotype | Sex | Age at onset | Age at diagnosis | Age at baseline | Race | Family history |
|---|---|---|---|---|---|---|
| **GP2 carriers** | | | | | | |
| PD | Female | 51-55 | 51-55 | NR | White | Yes |
| PD | Female | 46-50 | 46-50 | NR | Asian | NR |
| PD | Male | NR | 51-55 | 56-60 | Asian | No |
| Control | Female | NA | NA | 61-65 | White | NA |
| **VetPD carriers** | | | | | | |
| PD | Male | 61-65 | 66-70 | 76-80 | Native American | No |
| PD | Male | 76-80 | 76-80 | 76-80 | Native American | No |
| PD | Male | 61-65 | 61-65 | 66-70 | Asian | No |

NA = not applicable; NR = not reported

**Supplementary Table 4. Demographics of the PD participants screened for the VPS35 p.A320V mutation**

| **GP2 cohort** | **Early onset PD*** **(AAO < 50)** | **Late onset PD*** **(AAO >= 50)** | **Combined*** |
|---|---|---|---|
| n | 5,940 | 9,777 | 21,206 |
| Sex (male, %) | 61.2 | 61.5 | 60.7 |
| Age, average (SD; Min-Max) | 52.3 (9.7; 16-93) | 68.8 (8.4; 26-101) | 63.4 (11.6; 9-101) |
| AAO, average (SD; Min-Max) | 40.5 (7; 3-50) | 62.4 (8.3; 50-98) | 54.1 (13.2; 3-98) |
| Family history PD (%) | 27.5 | 42.58 | 30.1 |
| **VetPD cohort** | **Early onset PD*** **(AAO < 50)** | **Late onset PD*** **(AAO >= 50)** | **Combined*** |
| n | 193 | 1,219 | 1,413 |
| Sex (male, %) | 86.6 | 95.4 | 94.2 |
| Age, average (SD; Min-Max) | 60.8 (12; 21-87) | 74.65 (6.7; 50-93) | 72.7 (9; 21-93) |
| AAO, average (SD; Min-Max) | 40.3 (8.4; 12-49) | 66.04 (8; 50-88) | 62.5 (12; 12-88) |
| Family history PD (%) | 11.9 | 10.7 | 10.8 |
| **Mainland Spain cohort** | **Early onset PD (AAO < 50)** | **Late onset PD (AAO >= 50)** | **Combined*** |
| n | 90 | 477 | 567 |
| Sex (male, %) | 55.6 | 58.2 | 57.9 |
| Age, average (SD; Min-Max) | 62.9 (10.9; 36-84) | 78.8 (8.2; 56-102) | 76.23 (10.4; 36-102) |
| AAO, average (SD; Min-Max) | 42.1 (6.6; 16-49) | 62.90 (8; 50-84) | 59.60 (10.9; 16-84) |
| Family history PD (%) | 44.9 | 23.2 | 26.6 |
| **PDGenCI cohort** | **Early onset PD (AAO < 50)** | **Late onset PD (AAO >= 50)** | **All** |
| n | 23 | 118 | 141 |
| Sex (male, %) | 73.9 | 59.3 | 61.7 |
| Age, average (SD; Min-Max) | 54.8 (10.9; 37-76) | 70.70 (7.5; 53-86) | 68.1 (10; 37-86) |
| AAO, average (SD; Min-Max) | 39.7 (11.2; 9-49) | 63.96 (7.9; 51-83) | 59.99 (12.4; 9-83) |
| Family history PD (%) | 30.4 | 30.2 | 30.2 |

AAO = age at onset; GP2 = Global Parkinson's Genetics Program; Min = minimum; Max = maximum; PDGenCI = Parkinson's disease in the Canary Islands; SD = standard deviation; VetPD = Veterans Parkinson's Disease Genetics Initiative; *5,489 PD subjects from the GP2 cohort and 1 PD subject from the VetPD cohort did not have available age at onset, respectively. They are included in the combined category, but not in counts by AAO.

**Supplementary Table 5. Demographics of the controls screened for the VPS35 p.A320V mutation**

| | |
|---|---|
| **GP2 cohort** | |
| n | 9,172 |
| Sex (male, %) | 48.8 |
| Age, average (SD; min-max) | 62.7 (11.8; 1.6-104) |
| Family history PD (%) | 3.5 |
| **PDGenCI cohort** | |
| n | 63 |
| Sex (male, %) | 41.3 |
| Age, average (SD; min-max) | 62.8 (10.4; 36-86) |
| Family history PD (%) | 0 |

GP2 = Global Parkinson's Genetics Program; max = maximum; min = minimum; PDGenCI = Parkinson's disease in the Canary Islands; SD = standard deviation.

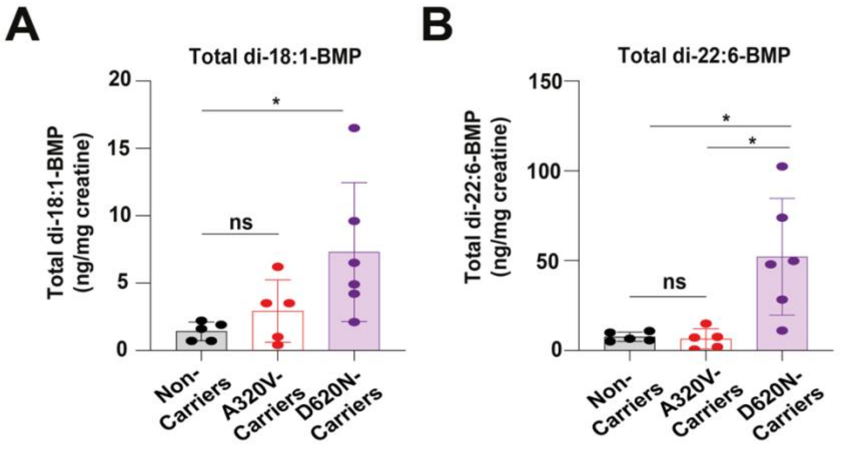

**Supplementary Figure 1.** Urinary bis(monoacylglycero)phosphate (BMP) level comparison across non-carriers, VPS35 p.A320V, and p.D620N carriers. Statistically significant differences between groups were assessed with Kruskal–Wallis test. Post-hoc Dunn's multiple comparison test was employed (ns = not significant; * $p < 0.05$.)

### Vet-PD Banner Authors

| <u>**Name**</u> | <u>**Institution**</u> |
|---|---|
| Sarah Pirio-Richardson, MD | Albuquerque VAMC, NM |
| Dana Sugar, MD | Albuquerque VAMC, NM |
| Marian Evatt, MD | Atlanta VAMC, GA |
| Jeanne Feuerstein, MD | Aurora VAMC, CO |
| Melissa Nirenberg, MD, PhD | Bronx VAMC, NY |
| Ruth Walker, MD, PhD | Bronx VAMC, NY |
| Vanessa Hinson, MD, PhD | Charleston VAHCS, SC |
| Christine Cooper, MD | Charleston VAHCS, SC |
| Gonzalo Revuelta, DO | Charleston VAHCS, SC |
| Brandon Barton, MD | Jesse Brown VAMC, Chicago, IL |
| Roshni Patel, MD | Jesse Brown VAMC, Chicago, IL |
| Roneil Malkani, MD | Jesse Brown VAMC, Chicago, IL |
| Karin Mente, MD | Cleveland VAHCS, OH |
| Aasef Shaikh, MD | Cleveland VAHCS, OH |
| Meagen Salinas, MD | Dallas VAHCS, TX |
| Juliana Atem, DNP | Dallas VAHCS, TX |
| Chris Hess, MD | Gainesville VAHCS, FL |
| Kalea Colletta, DO | Hines VAHCS, IL |
| Sandra Kletzel, PhD | Hines VAHCS, IL |
| Stuart Pang, MD | Honolulu VAMC, HI |
| George Jackson, MD, PhD | Houston VAMC, TX |
| Fariha Jamal, MD | Houston VAMC, TX |
| Vikas Singh, MD | Kansas City VAMC, MO |
| Dorothee Cole, MD | Loma Linda VAHCS, CA |

| | |
|---|---|
| Bradley Cole, MD | Loma Linda VAHCS, CA |
| Ronald Fernando, MD | Loma Linda VAHCS, CA |
| Adrienne Keener, MD | West Los Angeles VAHCS, CA |
| Sarah Perez, MD | New Orleans VAHCS, LA |
| Shannon Kilgore, MD | Palo Alto VAHCS, CA |
| James Morley, MD | Philadelphia VAMC, PA |
| John Duda, MD | Philadelphia VAMC, , PA |
| Gina Hopkins-Calligan, MD | Phoenix VAHCS, AZ |
| Steven Werder, DO | Phoenix VAHCS, AZ |
| Gregory Lazarz, MD | Phoenix VAHCS, AZ |
| Joseph Quinn, MD | Portland VAMC, OR |
| Kathryn Chung, MD | Portland VAMC, OR |
| Amie Hiller, MD | Portland VAMC, OR |
| Jessica Lehosit, DO | Richmond VAMC, VA |
| Kimmy Su, MD, PhD | Seattle VAMC, WA |
| Robert L. White, MD, PhD | St. Louis VAHCS, MO |
| Tanya Lin, MD | Tucson VAMC, AZ |

VAHCS = Veterans Affairs Healthcare System; VAMC = Veterans Affairs Medical Center
